# Characteristics of effective health education for older migrants from diverse cultural backgrounds – a scoping review protocol

**DOI:** 10.1101/2025.03.02.25322385

**Authors:** Cheng Yen Loo, Amy Page, Hazel Heng, Caroline Bulsara, Jacqueline Francis-Coad, Catherine M. Said, Adam Semciw, Ronald Shorr, Meg E. Morris, Anne-Marie Hill

## Abstract

**Objective:** The primary objective of this scoping review is to identify current evidence for the delivery of effective health education to older migrants from culturally diverse backgrounds. The secondary goals are to determine the characteristics of effective health education delivery for this population, how older migrants prefer to receive health education, and what cultural considerations influence the uptake of health education and put knowledge into practice.

**Introduction:** Access to health education is important to empower people to adopt healthy behaviours and to engage in informed decision-making about their well-being. Health education is not equally accessible in society and migrants who come from culturally diverse backgrounds can experience challenges in obtaining health information in a manner and format that is culturally responsive and linguistically appropriate. While many studies have reported the barriers and enablers to health information uptake among migrant communities, few have reported on what type of health education programs are most effective at imparting new skills, knowledge, and attitudes towards healthcare.

**Inclusion criteria:** Original qualitative, quantitative, and mixed methods published and unpublished studies that report on health education interventions to support and improve health education for migrants from culturally diverse backgrounds and their families will be eligible for inclusion. Studies that report on the type, format, and approaches older migrants prefer to access health education in either community or institutional settings will be included.

**Methods:** This scoping review will be conducted following the Joanna Briggs Institute’s method for evidence synthesis. In consultation with a research librarian, a literature search strategy will be developed comprising keywords, index terms, and medical subject headings. Electronic databases: PubMed, ProQuest Public Health, CINAHL, Embase, PsychINFO, and Web of Science will be searched for relevant studies with no date limitations. A Google Advanced and ProQuest Thesis and Dissertation searches will be conducted to capture grey literature. All references will be imported into Covidence® where two independent reviewers will perform study selection and data extraction. Key concepts and evidence will be presented through a narrative summary of findings, which will include the identification of findings that align with the scope of this review, an overview of the research evidence, and the identification of research gaps.

## INTRODUCTION

Health education is an important component of effective health care, contributing to positive health behaviours and consequent improvements in psycho-social aspects of health (1). Despite the demonstrated benefits of providing health education, a lack of health education is a widespread problem among the general population in many countries (1). Furthermore, migrants face additional barriers accessing health education due to a range of factors such as having limited language proficiency (2), poor digital literacy (3), limited social support networks (4) and different socio-cultural values than the dominant population in their host country (5). A recent scoping review on the effectiveness of patient education for migrants with heart disease, found significant gaps in reporting the adaptation process for educational interventions for migrants from culturally diverse backgrounds (6). It is important to recognise that delivering trusted, and effective health education involves more than having access to websites and pamphlets translated into different languages (7). Health education appears to be most effective when information is delivered in negotiated partnership with migrant communities that empower them to make informed lifestyle and behavioural changes in a culturally appropriate and safe manner (8-10).

Cultural competency requires health educators to learn about the particular norms, behaviours and practices of how other cultures prefer to engage in cross-cultural dialogue (11). It also requires health educators to engage in flexible thinking to adjust professional work styles to meet the values, expectations, and preferences of culturally diverse migrant groups (11, 12). An extension of delivering culturally competent health education is to do so in a culturally safe manner, which requires health educators to acknowledge and respect the cultural beliefs and values of migrant communities, recognise homeland practices and any historical trauma associated with their migrant experience (13-15).

In countries with a large ageing migrant population, delivering health education focused on healthy ageing in a culturally competent and safe manner is of growing importance (14). Yet, a recent study among older migrants found that they were less likely to engage in help-seeking behaviour from mainstream channels, instead preferring to draw upon alternative pathways rooted in their health beliefs shaped by cultural and religious context (16). For countries such as Australia, this presents a serious public health concern, given that a recent national ageing research report (17) found that Australia’s European-born population aged 65 years and over is expected to decline over the next decade while the Asian-born population in Australia aged 65 years and over is projected to reach 1.5 million by 2056 (17). According to the 2016 Australian Bureau of Statistics census (18), almost 40% of all migrants from culturally diverse backgrounds were aged 50 years and over compared to 32.4% of Australia’s total population aged 50 and over.

### Cultural beliefs and help-seeking behaviour

Older migrants from culturally diverse backgrounds are more suspectable to poorer health outcomes, in part because they are less likely to undertake routine health checks through approved mainstream channels (19, 20). This was highlighted in research that found migrant communities in Australia exhibited poorer uptake of mental health services compared to the general population, despite reporting higher rates of disability attributed to psychological distress, particularly among refugees and asylum seekers (19, 21).

A scoping review reported that migrants from culturally diverse backgrounds may have different beliefs about illnesses and treatments to that of health professionals, depending on their culture and religious beliefs (22). Some cultural groups under-utilised and even abstained from seeking treatment for certain health conditions due to societal and cultural stigma (20). Chinese-Australian migrants reported a higher preference to obtain mental health support from informal channels such as family to avoid shame (23). In a separate study from conducted in the United Kingdom (UK), it was reported that certain practices such as religious fasting modified how migrants from South Asia and the Middle East regions took their medication; sometimes opting to cease treatment altogether against doctors’ advice (22, 24). A complementary study that explored the heal practices of older Ghanaian migrants residing in the UK, reported a strong preference to use herbal medicine, faith-based healing, and traditional healers to address ailments rather than seek council from a doctor (16).

### Accessing trusted health education

Although an abundance of health information is available in most countries with developed health systems, awareness of and access to trusted health information (e.g: government-approved information) remains disproportionately low in some migrant communities. A recent systematic review found that some migrant groups were more reliant on diaspora media for obtaining health education during the COVID-19 pandemic due to limited access to approved health information in their preferred language (25). Compounding this difficulty was the phenomenon called infodemic – where torrents of online information containing either false or misleading information flood uncredited social media outlets leading to the production of misinformation and disinformation (25, 26). The spread of misinformation and disinformation can undermine trust and create public doubt about where to source health education (25).

Many studies have reported on the barriers and enablers experienced by older migrants from culturally diverse backgrounds accessing health education (27-29). However, no reviews have been undertaken to examine the breadth and depth of evidence available in this area.

There is a need to map the current evidence available to examine the key factors that make health education effective for older migrants from culturally diverse backgrounds, including their preferred method of receiving education, the cultural factors that influence their engagement, and how those considerations intersect to foster cultural safety. A scoping review can mitigate this gap by identifying the breadth of available evidence on delivering health education to older migrants from culturally diverse backgrounds (30, 31). Conducting a scoping review will also clarify the concepts related to the subject field and identify the contextual factors that inform recommendations (32).

The aim of this scoping review is to identify current evidence for the delivery of effective health education to older migrants from culturally diverse backgrounds.

## REVIEW QUESTION

### Primary Question

What are the characteristics of effective health education for older migrants from culturally diverse backgrounds?

### Secondary question (i)

How do older migrants from culturally diverse backgrounds prefer to receive health education?

### Secondary question (ii)

What cultural considerations influence older migrants from culturally diverse backgrounds to accept and engage in the recommendations provided in health education?

### Secondary question (iii)

What are the attributes of an education program for older migrants from culturally diverse backgrounds that make it culturally safe?

## INCLUSION CRITERIA

The review will use the Population, Concept and Context (PCC) framework as recommended in the Joanna Briggs Institute Manual for Evidence Synthesis (33).

### Population

Studies conducted in populations that specifically focus on older migrants from culturally diverse backgrounds will be eligible for inclusion. The population of older migrants is defined as any foreign-born person aged 65 years and over who moved across international borders temporarily or permanently and resides in a nation that speaks English as the official language of government, and whose dominant population shares similar customs, values and cultural heritage (34). It is expected the study population will have distinctive customs, history, or culture unique from a dominant population of Anglo-Celtic heritage.

### Concept

This scoping review will search for studies that evaluate health education interventions for older migrants from culturally diverse backgrounds, living in countries that have substantial cohorts of overseas-born older adults. Preliminary searches suggest that studies conducted in Australia and other English-speaking countries such as New Zealand, the UK, Canada, and the United States (US) will be identified. Other concepts are studies that report on effective characteristics of delivering health education to older migrants of culturally diverse backgrounds, their preferred mode of receiving health education, the cultural considerations that can influence the uptake and practice of health knowledge, and attributes of such interventions that deem it culturally safe.

### Context

This scoping review will include studies published in English. Both community-based and institutional (i.e. hospital and residential aged care) health education studies designed for migrant communities will be included. Studies will be geographically limited to English-speaking nations with large culturally diverse migrant populations to focus on generalisation to the topic of these health care systems. These countries include Australia, Canada, New Zealand, the UK, and the US.

### Type of Sources

This scoping review will consider experimental, quasi-experimental study designs including randomised controlled trials, non-randomised controlled trials, before and after studies and interrupted time-series studies. In addition, analytical observational studies including prospective and retrospective cohort studies, case-controlled studies and analytical cross-sectional studies will also be considered for inclusion. Case series, individual case reports and conference abstracts will be excluded. Qualitative studies may be considered that focus on qualitative data including but not limited to, designs such as phenomenology, grounded theory, ethnography, qualitative description, action research and feminist research.

## METHODS

The proposed scoping review will be conducted following the Joanna Briggs Institute’s methodology for scoping reviews (31, 33). This scoping review is registered in Open Science Framework (https://osf.io/5bu3e/).

Because this is an under-researched field of study with no previous reviews focusing on older migrant populations, using a scoping review design was deemed appropriate for generating an overview of available literature that has been conducted on a specific topic and answering broad questions about the characteristics of such education (33). Based on the preliminary database search results, it is anticipated there will not be a large quantity of relevant literature that reports on effective health education for older migrants of culturally diverse backgrounds. This scoping review will use the Preferred Reporting Items for Systematic Reviews and Meta-Analyses extension for Scoping Review (PRISMA-ScR) checklist to guide the methodology of the review and the reporting of the results (30).

### Search Strategy

The search strategy will aim to locate both published and unpublished studies. A three-step search strategy will be utilised in this review. First, an initial limited search of MEDLINE (PubMed) and ProQuest will be undertaken to identify articles on the topic. The text words contained in the titles and abstracts of relevant articles, and the index terms used to describe the articles will then be used to develop a full search strategy for all relevant databases/information sources (Appendix 1). The search strategy, including all identified keywords, MeSH, and index terms (Table 1), will be adapted for MEDLINE, CINAHL, Embase, and Web of Science. For grey literature, Google Advanced Search and ProQuest Dissertations and Thesis will be used to search for reports, studies, and programs published by government, non-profit, and educational institutions.

**Table 1:** Topic Facet and MeSH terms

| **Topic Facet** | **Index terms or MeSH Terms** |
| --- | --- |
| **Migrant** | “Migrant” OR “CaLD” OR “Culturally and Linguistically Diverse” OR “Non-English Speaking” OR “Immigrant” OR “Ethnic Minority” OR “Multicultural” |
| **Older Adult** | “Older Adult” OR “Elder*” OR “Geriatric” OR “Older person” OR (MH “aging”) OR (MH “aged”) OR “Seniors” |
| **Health Education** | “Health Education” OR “Health Promotion” OR “Public Health Awareness” OR “Health Literacy” OR “Health Translation” OR “Health Awareness” OR “Patient Education” |

Only studies published in English from inception to the present will be included because the review will focus on migrant populations residing in countries that speak English as the national language.

### Study/Source of Evidence Selection

Following the search, all the citations will be collated and uploaded into Covidence® and duplicates will be removed. Following a pilot test, titles and abstracts will then be screened by two independent reviewers against the inclusion criteria.

In the second screening level, the authors will again use the inclusion and exclusion criteria to complete a full-text review. To ensure all relevant studies are captured during the full-text review, citation chaining will be adopted to identify articles that meet the inclusion criteria not captured during the database searches (35). Studies excluded during this screening phase against the inclusion criteria will be documented in the scoping review. Any disagreements between the reviewers will be resolved through discussions involving a third researcher or with additional notes written against the relevant citation. The results from the search and the study inclusion process will be reported in full in the final scoping review and presented in a PRISMA diagram (30).

### Data Extraction

Data will be extracted from research articles included in the scoping review independently by two reviewers using a basic data extraction tool in MS Excel (Appendix 2). The data to be extracted will include specific details of the publication, subject population, concept, context, participant characteristics, research design and key findings relevant to the review questions. If any additional information is required to clarify doubts about some of the study’s information, the authors of the evidence sources will be contacted by the reviewers.

To determine the cultural competency of each study and the degree to which cultural safety was integrated into the development of each intervention or health education program, the research articles will be evaluated against the 10 domains of the culturagram (Appendix 3). The culturagram is an assessment tool designed to systematically gather in-depth information on how a person’s culture affects their perspectives of life and their situation (36, 37).

Originating in social work research, they were designed to help caseworkers assess the worldview of migrant clients/patients more effectively so that culturally appropriate and accessible interventions could be developed (38). The culturagram is underpinned by the notion that migrant groups are not homogenous and the risk of over-generalising their characteristics may lead to incorrect stereotypes (37, 39).

The New World Kirkpatrick Model (40) for evaluating education and training programs will be used to appraise the impact of each intervention for older migrants of culturally diverse backgrounds. The Kirkpatrick Model is a versatile framework used to evaluate short to long-term outcomes of training programs and individual and group behaviours (40). It has been used in a range of research fields including organisational studies (41), hospital studies (42), and simulation training (43). In this scoping review, the Kirkpatrick Model will be used to evaluate the impact of each intervention/health education program according to the reported outcomes which will then be assigned to one of the four evaluation levels (Appendix 4):

- Level 1(Reaction): Client/patient level of satisfaction, engagement and perceived value of the intervention/education program;
- Level 2 (Learning): The level of change in client/patient understanding of health education before and after the program;
- Level 3 (Behaviour): The level of behavioural change attributed to the intervention/health education program; and
- Level 4 (Results): The overall impact of the training to the individual or family or migrant community.

Following Joanna Briggs Institute’s recommendations for conducting scoping reviews, critical appraisal of included studies or reports will not be undertaken. Since the review will seek to comprehensively map the body of literature around education for older migrants from culturally diverse backgrounds, methodological quality will not be used to determine inclusion/exclusion from the review (33).

### Data Analysis and Presentation

The extracted data will be presented in tabular format and will report the distribution of studies by study design, participants (sample size and population), location, aims, intervention, evaluation strategy, and outcomes. Qualitative content analysis (44, 45) taking a deductive approach based on the cultural safety and education framework will be used to categorise and interpret textual data to enable the identification of gaps, patterns, and insights that inform the broader research question. We will use descriptive statistics (e.g. frequencies, means, percentages) to report on the cultural competency of each study. The efficacy of each study appraised against the Kirkpatrick model will be presented in tabular format and as narrative summaries that align with the aim of this scoping review.

## AUTHOR CONTRIBUTIONS

Conceptualisation, CYL, A-MH, HH, AP, CB, JF-C, MM; Formal analysis, CYL, A-MH, HH, AP, CB; CS, AS, RS; Funding acquisition, A-MH, MM; AP, HH, CB, JF-C, CS, AS, RS; Investigation, CYL, A-MH, AP, HH, CB, JF-C, MM; Methodology, CYL, A-MH, HH, AP, CB, JF-C, MM; Project Administration, CYL, A-MH, HH; AP, Writing – original draft, CYL, A-MH, HH; Writing – review and editing, CYL, A-MH, HH, AP, CB, JF-C, MM. All authors have reviewed and agreed to the final submitted version of the protocol.

## ETHICS STATEMENT

No Ethics Committee was needed for this study.

## Data Availability

All data produced in the present work are contained in the manuscript

## Appendix 1 Search Strategy

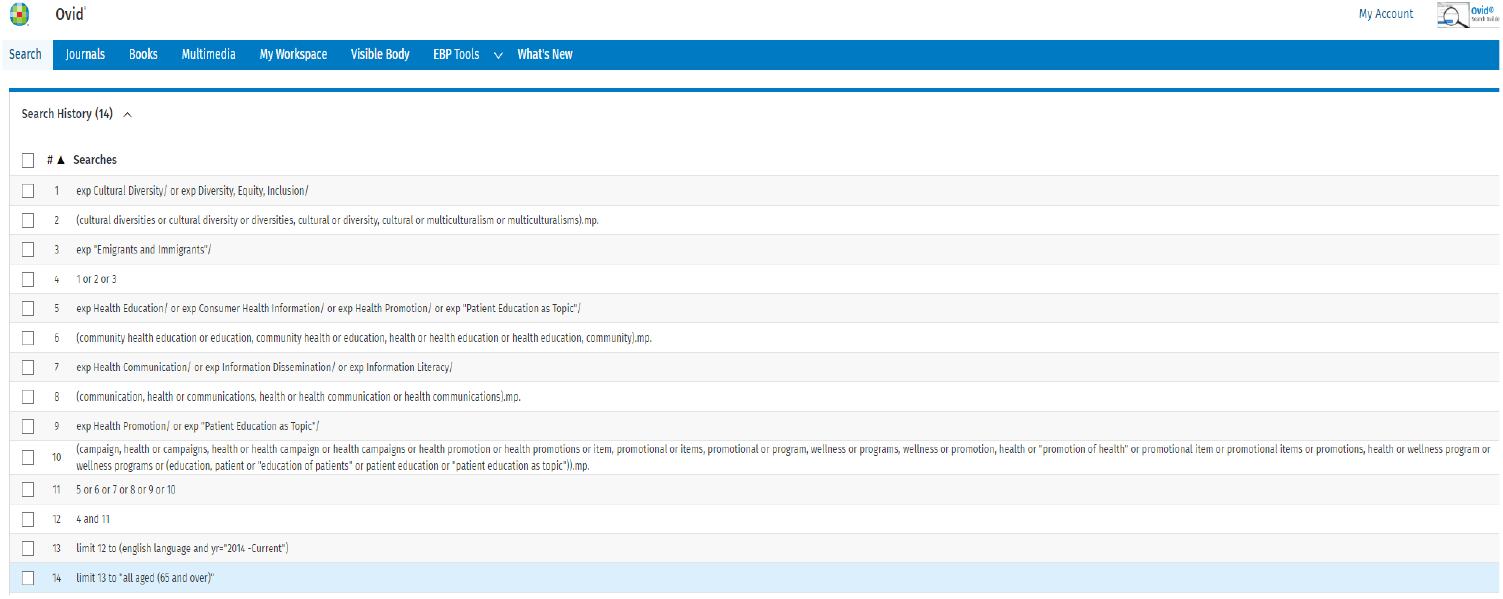

## Appendix 2 Baseline Data Extraction Tool

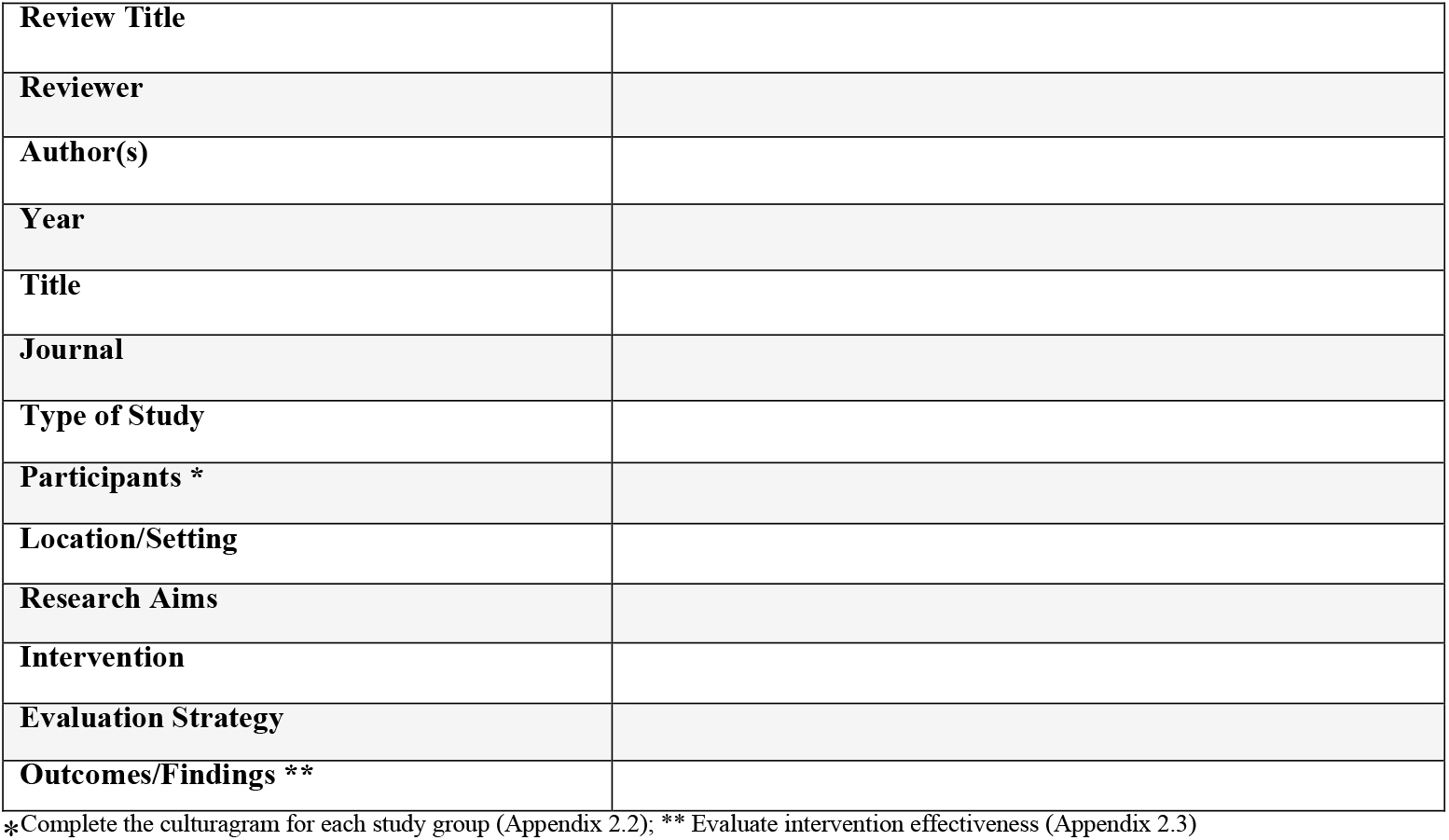

## Appendix 3 Culturagram Evaluation Tool

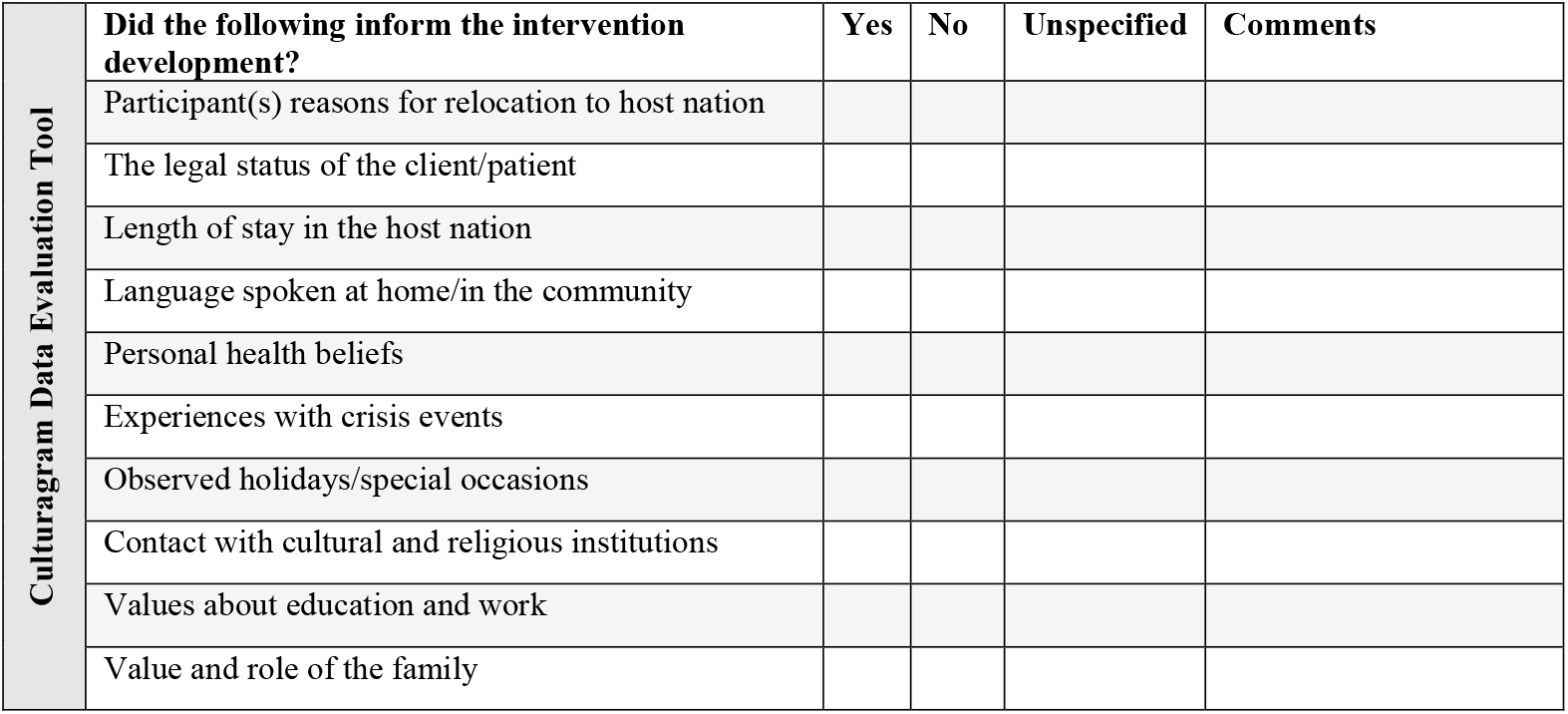

## Appendix 4 Intervention Effectiveness Evaluation Tool (New World Kirkpatrick Model)

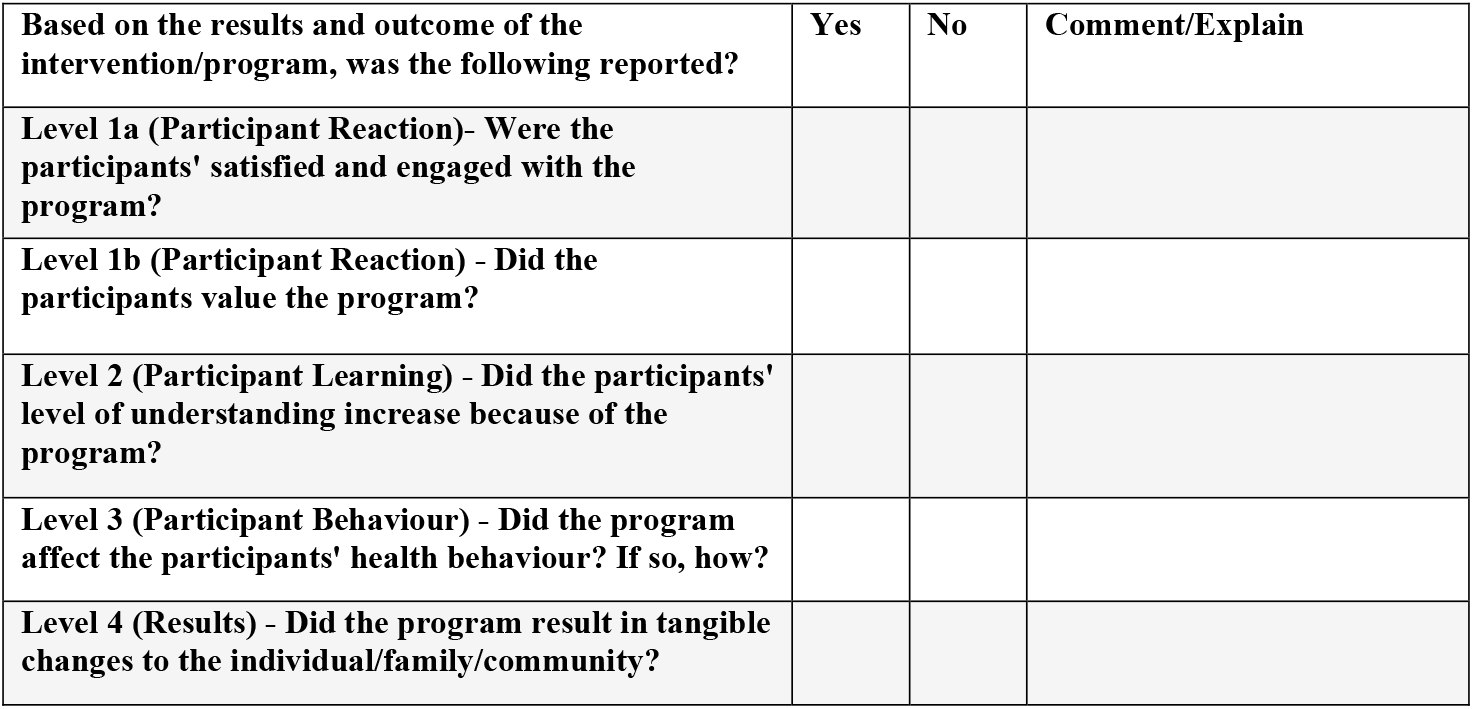

## REFERENCES

1. Rizvi DS. Health education and global health: Practices, applications, and future research. J Educ Health Promot. 2022;11:262.

2. Al Shamsi H, Almutairi AG, Al Mashrafi S, Al Kalbani T. Implications of Language Barriers for Healthcare: A Systematic Review. Oman Med J. 2020;35(2):e122.

3. Fitzpatrick PJ. Improving health literacy using the power of digital communications to achieve better health outcomes for patients and practitioners. Front Digit Health. 2023;5:1264780.

4. Pan H, Qualter P, Barreto M, Stegen H, Dury S. Loneliness in Older Migrants: Exploring the Role of Cultural Differences in Their Loneliness Experience. Int J Environ Res Public Health. 2023;20(4).

5. Khatri RB, Assefa Y. Access to health services among culturally and linguistically diverse populations in the Australian universal health care system: issues and challenges. BMC Public Health. 2022;22(1):880.

6. Zhang L, Ghisi GLM, Shi W, Pakosh M, Main E, Gallagher R. Patient education in ethnic minority and migrant patients with heart disease: A scoping review. Patient Educ Couns. 2025;130:108480.

7. Migrant and Refugee Women’s Health Partnership. Enhancing health literacy strategies in the settlement of migrant and refugee women. Canberra: Migrant and Refugee Women’s Health Partnership,; 2018.

8. Hardy BJ, Filipenko S, Smylie D, Ziegler C, Smylie J. Systematic review of Indigenous cultural safety training interventions for healthcare professionals in Australia, Canada, New Zealand and the United States. BMJ Open. 2023;13(10):e073320.

9. Pocock NS, Chan Z, Loganathan T, Suphanchaimat R, Kosiyaporn H, Allotey P, et al. Moving towards culturally competent health systems for migrants? Applying systems thinking in a qualitative study in Malaysia and Thailand. PLoS One. 2020;15(4):e0231154.

10. Goodman C, Lambert K. Scoping review of the preferences of older adults for patient education materials. Patient Educ Couns. 2023;108:107591.

11. Garran AM, Werkmeister Rozas L. Cultural Competence Revisited. Journal of Ethnic & Cultural Diversity in Social Work. 2013;22(2):97–111.

12. Ogbogu PU, Noroski LM, Arcoleo K, Reese BD, Apter AJ. Methods for Cross-Cultural Communication in Clinic Encounters. The Journal of Allergy and Clinical Immunology: In Practice. 2022;10(4):893–900.

13. Smye Victoria, Josewski Viviane, Kendall Emma. Cultural Safety: An Overview. Canada: Mental health Commission of Canada; 2010.

14. Kurtz DLM, Janke R, Vinek J, Wells T, Hutchinson P, Froste A. Health Sciences cultural safety education in Australia, Canada, New Zealand, and the United States: a literature review. Int J Med Educ. 2018;9:271–85.

15. Polaschek. Cultural safety: a new concept in nursing people of different ethnicities. Journal of Advanced Nursing. 1998;27(3):452–7.

16. Krzyż EZ, Lin H-R. Health-seeking behavior in older immigrants: a concept analysis. Educational Gerontology. 2024;50(5):386–401.

17. Centre of Excellence in Population Ageing Research. Migration and ageing: How cultural and linguistic diversity is set to boom among older Australians. Canberra: Australian Research Council, ; 2024.

18. Cultural Diversity in Australia, 2016 [Internet]. Australian Bureau of Statistics,. 2016 [cited 5 January 2025].

19. Teo K, Churchill R, Riadi I, Kervin L, Wister AV, Cosco TD. Help-Seeking Behaviors Among Older Adults: A Scoping Review. Journal of Applied Gerontology. 2022;41(5):1500–10.

20. Federation of Ethnic Communities Councils of Australia. Review of Australian Research on Older People from Culturally and Linguistically Diverse Backgrounds. Canberra: Federation of Ethnic Communities Councils of Australia,; 2015.

21. Slewa-Younan S, Krstanoska-Blazeska K. A Literature Review on Mental Health Stigma in Three Spcific Cultrually and Linguistically Diverse Communities: Arabic, African and Chinese Sydney Western Sydney University 2022.

22. Chauhan A, Walton M, Manias E, Walpola RL, Seale H, Latanik M, et al. The safety of health care for ethnic minority patients: a systematic review. Int J Equity Health. 2020;19(1):118.

23. Shi W, Shen Z, Wang S, Hall BJ. Barriers to Professional Mental Health Help-Seeking Among Chinese Adults: A Systematic Review. Front Psychiatry. 2020;11:442.

24. Alhomoud F, Dhillon S, Aslanpour Z, Smith F. South Asian and Middle Eastern patients’ perspectives on medicine-related problems in the United Kingdom. Int J Clin Pharm. 2015;37(4):607–15.

25. Goldsmith LP, Rowland-Pomp M, Hanson K, Deal A, Crawshaw AF, Hayward SE, et al. Use of social media platforms by migrant and ethnic minority populations during the COVID-19 pandemic: a systematic review. BMJ Open. 2022;12(11):e061896.

26. Borges do Nascimento IJ, Pizarro AB, Almeida JM, Azzopardi-Muscat N, Gonçalves MA, Björklund M, et al. Infodemics and health misinformation: a systematic review of reviews. Bull World Health Organ. 2022;100(9):544–61.

27. Abba-Aji M, Stuckler D, Galea S, McKee M. Ethnic/racial minorities’ and migrants’ access to COVID-19 vaccines: A systematic review of barriers and facilitators. J Migr Health. 2022;5:100086.

28. Magee L, Knights F, McKechnie DGJ, Al-Bedaery R, Razai MS. Facilitators and barriers to COVID-19 vaccination uptake among ethnic minorities: A qualitative study in primary care. PLoS One. 2022;17(7):e0270504.

29. Commonwealth Department of Health. Actions to support older Culturally and Linguistically Diverse people: A guide for aged care providers. Canberra: Commonwealth Department of Health; 2019.

30. Tricco AC, Lillie E, Zarin W, O’Brien KK, Colquhoun H, Levac D, et al. PRISMA Extension for Scoping Reviews (PRISMA-ScR): Checklist and Explanation. Annals of Internal Medicine. 2018;169(7):467–73.

31. Peters MDJ, Marnie C, Tricco AC, Pollock D, Munn Z, Alexander L, et al. Updated methodological guidance for the conduct of scoping reviews. JBI Evid Synth. 2020;18(10):2119–26.

32. Pollock D, Evans C, Menghao Jia R, Alexander L, Pieper D, Brandão de Moraes É, et al. “How-to”: scoping review? Journal of Clinical Epidemiology. 2024;176:111572.

33. Aromataris E, Lockwood C, Porritt K, Pilla B, Jordan Z. JBI Manual for Evidence Synthesis. 2024.

34. Wellings B, Mycock A. The Anglosphere: Continuity, Dissonance and Location: British Academy; 2019.

35. Hirt J, Nordhausen T, Appenzeller-Herzog C, Ewald H. Citation tracking for systematic literature searching: A scoping review. Research Synthesis Methods. 2023;14(3):563–79.

36. Congress E, Kurnick K. The culturagram matrix: Domains of migration identities. In: Rich GJ, Kuriansky J, Gielen UP, Kaplin D, editors. Psychosocial Experiences and Adjustment of Migrants : Coming to the USA. Chantilly, UNITED STATES: Elsevier Science & Technology; 2023. p. 111–24.

37. Congress E, Kung W. Using the Culturagram and an Intersectional Approach in Practice With Culturally Diverse Families. In: Congress E, Gonzalez M, editors. Multicultural Perspectives in Working With Families. 4 ed. New York: Springer Publishing Company; 2020. p. 3–25.

38. Jani J, Okundaye J. Teaching Note: The Culturagram: An Educational Tool to Enhance Practice Competence With Diverse Populations. Journal of Baccalaureate Social Work. 2014;19:53–63.

39. Chau RCM, Yu SWK, Tran CTL. Understanding the diverse health needs of Chinese people in Britain and developing cultural sensitive services. Journal of Social Work. 2011;12(4):385–403.

40. Kirkpatrick JD, Kirkpatrick WK. Kirkpatrick’s Four Levels of Training Evaluation. Alexandria, United States: American Society for Training & Development; 2016.

41. Paull M, Whitsed C, Girardi A. Applying the Kirkpatrick model: Evaluating an “interaction for learning framework” curriculum intervention. Issues in Educational Research. 2016;26(3):490–507.

42. Francis-Coad J, Farlie M, Haines T, Black L, Weselman T, Cummings P, et al. Revising and evaluating falls prevention education for older adults in hospital. Health Education Journal. 2023;82(8):878–91.

43. Chia N-H, Cheung VK-L, Lam ML-Y, Cheung IW-K, Wong TK-Y, So S-S, et al. Harnessing power of simulation training effectiveness with Kirkpatrick model in emergency surgical airway procedures. Heliyon. 2022;8(10).

44. Assarroudi A, Heshmati Nabavi F, Armat MR, Ebadi A, Vaismoradi M. Directed qualitative content analysis: the description and elaboration of its underpinning methods and data analysis process. Journal of Research in Nursing. 2018;23(1):42–55.

45. Kleinheksel AJ, Rockich-Winston N, Tawfik H, Wyatt TR. Demystifying Content Analysis. American Journal of Pharmaceutical Education. 2020;84(1):7113.

